# “Effectiveness of Sexual Health Education Interventions Addressing Sexual Violence and Sexual Safety for Adolescents: A Systematic Review Protocol”

**DOI:** 10.1101/2025.06.20.25328154

**Authors:** Margaret Noonan, Patrick Cotter, Josephine Hegarty, James O Mahony, Sharon Lambert

## Abstract

**Background:** Sexual health education interventions should provide adolescents with the knowledge and skills to assist them to navigate and develop safe and healthy relationships allowing them to positively transition from adolescence to adulthood. However, many sexual health education interventions omit addressing sexual violence and sexual safety. Of the interventions that do address sexual violence and sexual safety many are poorly or not at all evaluated. This paper details a protocol for a mixed-methods systematic review to critically appraise the evidence on the effectiveness of sexual health interventions that address sexual violence and sexual safety for adolescents.

**Methods:** A systematic review of the literature following the Preferred Reporting Guidelines for Systematic Reviews and Meta-analyses (PRISMA) reporting guidelines will be undertaken. A search of the following databases will be conducted: CINAHL, PSYCHINFO, ERIC, EMBASE, PubMed, JSTOR, Scopus, PyscArticles, Soc Index with Full Text, Humanities Full Text (H. W. Wilson) and the British Education Index. Citations will be evaluated against predefined eligibility criteria using a two-stage screening process which will be conducted independently in pairs. Data will be extracted from eligible papers using pilot-tested data extraction forms focusing on measurements of effectiveness of the intervention and the impact of the intervention. Quality of evidence will be assessed using the Mixed Methods Appraisal Tool (MMAT). The Template for intervention and description (TIDieR)) framework will be utilised to describe the included interventions.

**Discussion:** This study will describe sexual health education interventions that address sexual violence and sexual safety for adolescents and their efficacy. The evidence will help inform future planning for sexual health education interventions which address sexual violence and/or safety. OSF Registration: https://doi.org/10.17605/OSF.IO/YP2XQ

## Introduction

Sexual violence is a worldwide public health problem that affects every gender, sexuality, and society (1). The World Health Organisation estimates that 1 in 5 (18%) of girls have experienced child sexual abuse and while acknowledging the magnitude of child sexual abuse affecting boys has yet to be uncovered, with estimates that 1 in 7 boys will experience some form of sexual violence (2). Banvard et al. (2020) (3) argue that preventing adolescent sexual violence could significantly improve long-term health outcomes for this population, while also having a substantial impact on public health and health economics.

Lameiras-Fernández et al (2021) (4 pg2) define sexual health education as “any combination of learning experiences aimed at facilitating voluntary behaviour conducive to sexual health”. Article 19 of the United Nations Convention on the Rights of the Child (UNCRC) (1989) obliges states to take appropriate measures to protect children from all forms of violence, (5) while the Council of Europe Convention, (Istanbul Convention, 2014), states that primary sexual violence prevention programs should be provided through educational programs, (6). In 2007, The Lanzarote Convention specified that children need to be taught about the risks of sexual exploitation, sexual abuse and how to stay safe (7). The United Nations Educational, Scientific, and Cultural Organisation (UNESCO) suggests that, comprehensive sexuality education plays a central role in the preparing young people for a safe productive fulfilling life, (8).

Studies have shown that students who received education about sexual violence before attending college had less rape myth acceptance when in college (9) and have less chance of being a victim or a perpetrator (10). In a study by Denning et al (2023) (11) survivors of sexual violence were more than twice as likely to report never having sexual education or engaging in sex education for an insufficient duration compared to those who had never experienced sexual violence.

However, sex education programs have traditionally focused on abstinence or taken a female gender lens, all of which serve to alienate groups of students and fail to address key areas about sexual violence and safety (6,10,12,13). Excluding sexual violence from sexuality education may render young people vulnerable to victimization and perpetration and may deter them from seeking help if needed(14). UNESCO determine the key concepts of a comprehensive sex education program to include relationships; values; rights; culture and sexuality; understanding gender; violence and staying safe; skills for health and well-being; the human body and development; sexuality and sexual behaviour, and sexual and reproductive health. In one of their systematic reviews of sexual education interventions more than 30% of interventions only briefly cover sexual violence with 10% not covering sexual abuse/violence at all (8,15).

The UNESCO Global review of Sexuality Education (2020) (15) also highlights that many sexuality education interventions do not provide enough coverage for marginalised groups including those living with a disability, LGBTQ+ communities, and male students. Higher Education Institutions have developed policies and programs, but students in third level education believe that this is too late to receive this education and suggest that education should start much earlier in second level education and in schools ideally before student’s first romantic/sexual encounter (**10**).

### Review Aim

This systematic review aims to gather, assess, and synthesize evidence on the effectiveness of sexual health education interventions that address sexual violence and sexual safety among adolescents.

### Review Questions

1. What sexual health educational interventions for adolescents that specifically address sexual violence and sexual safety have been implemented and evaluated?
2. Within the included interventions, what content specifically related to sexual violence and sexual safety?
3. How was effectiveness of the intervention evaluated and what tools were used to measure effectiveness?
4. How effective were the interventions in addressing the primary outcomes of attitudes, beliefs, knowledge, intentions related to sexual violence and sexual safety and the secondary outcomes of behaviours related to sexual violence and sexual safety.

## Methods

### Protocol and Registration

This protocol is registered at Open Science Framework, under this number of identification: osf.10/854bv. The protocol was prepared and reported according to the “Preferred Reporting Items for Systematic Review and Meta-analysis Protocols” (16).

### Eligibility criteria

The eligibility criteria are organised according to the Population, Intervention, Comparison, Outcome, (PICO) framework (17).

### Types of Populations

The population of interest will be adolescents. The World Health Organisation defines adolescence as the period between childhood and adulthood, 10 years to 19 years of age (18). For this review, studies with participants aged from 10 years to 19 years in any pre-university setting will be considered for inclusion. Any studies aimed at a younger population, an older population, those in third level education or educators will be excluded.

### Types of Interventions or Exposure

The review will examine any sexual health educational interventions that include content specifically addressing sexual violence and/or sexual safety. The WHO defines sexual violence “as any act, attempt to obtain a sexual act, unwanted sexual comments or advances, or acts to traffic, or otherwise directed against a person’s sexuality using coercion, by any person, regardless of their relationship to the victim, in any setting, including but not limited to school and work”, (19).

Sexual safety is a broader concept than safe sex which is about protecting one’s health. Sexual safety includes the emotional, psychological and relational safety ensuring that all aspects of the sexual experience are secure and respectful (20). Sexual health education in the context of this review means education that equips adolescents with the knowledge, skills, attitudes and values that help them to protect their health, develop healthy respectful relationships, make responsible choices and understand their rights and those of others and to protect the rights of others (21). There will be no restriction on characteristics of the educational intervention such as mode of delivery, participant group size, setting of intervention, duration of intervention, or the developers and facilitators of interventions.

### Types of Studies

Empirical studies of any design provided they have a clear evaluative aim, and were conducted in any setting will be included. Relevant linked process evaluations will also be included. The timing of when measures are applied may also vary between studies with some studies completing pre-and post-test measures and others reporting only post-test measures. Systematic reviews will be tagged for reference list checks.

### Type of Outcomes

The primary outcomes are measures of effectiveness of sexual health education interventions that specifically address sexual violence and sexual safety, relating to the knowledge, attitudes, beliefs, intentions of adolescents towards sexual violence and sexual safety. Due to the nature of the interventions, there may be a variety of outcome measures used e.g. a rape myth acceptance scale to measure changes in attitudes and beliefs.

The secondary outcomes will be measures of intentions and behaviours of adolescents linked to sexual violence, sexual safety, and sexual health.

As the most recent Global Status review by UNESCO (15) of the comprehensive sexuality education was in 2020 a time limit of 10 years will apply. This time limit affords 5 years either side of the review therefore progress as per UNSECO recommendations should be visible in the later education programs.

### Search strategy

Potentially relevant citations will be identified by searching electronic databases. A comprehensive search strategy will be developed and implemented across databases to retrieve potentially relevant studies. Databases to be searched will include CINAHL, PSYCHINFO, ERIC, EMBASE, PubMed, JSTOR, Scopus, PyscArticles, Soc Index with Full Text, Humanities Full Text (H. W. Wilson) and Social Sciences Full Text (H. W. Wilson) and the British Education Index.

The reference list of included studies will be reviewed for further potentially relevant studies. When indicated, study authors will be contacted to obtain further information to clarify study characteristics. When study protocols are identified without sourcing subsequent publication of results, authors will be contacted to check if results have been published and determine eligibility for inclusion in this review.

### Study records

#### Search and Screening Process

The files from each database will be collected and imported into Covidence. Duplicates will be removed automatically within the software.

#### Selection Process

MN will assess the eligibility of all identified studies by reviewing titles and abstracts independently in pairs with PC, JH, JOM and SL. The reviewers will be blinded to each other’s decisions. A third reviewer will resolve conflicts. If titles and abstracts do not provide enough information to determine the eligibility of a particular study, the corresponding paper will be subjected to a full-text examination. Two reviewers will review all the full text papers and be blinded to each other’s decisions. A third reviewer will resolve any conflicts at either title/abstract screening or full text screening. The rationale for excluding studies from the review at the full text screening stage will be recorded and reported in the systematic review. To illustrate the study selection process and reasons for exclusion, a PRISMA(22) flow chart will be used to visually represent the process.

### Data extraction

The process of data extraction will begin with collecting key details including the authors, title, year of publication, language, country of publication, from each included paper.

Next, the study characteristics will be outlined, covering aspects such as the study location and design, details of the facilitators who conducted and delivered the intervention, and the number of students involved. Additional demographic information such as the age, race, and gender of the participants will also be recorded.

The characteristics of the intervention will be examined using the Template for Intervention Description and Replication Checklist (TIDieR) framework (23), ensuring a systematic approach to understanding its components and implementation.

For the outcome measures, an extraction tool will be piloted on a subset of the studies to confirm that the necessary data to answer the review question is being accurately collected. Depending on the findings from the pilot, the data extraction tool may be refined to capture the most relevant information, facilitating a meaningful synthesis of the results.

### Quality Assessment

All included studies will be subject to quality assessment and a risk of bias assessment. Several intervention types will be used in this systematic review; therefore, quality assessment will be carried out using the Mixed Methods Appraisal Tool (MMAT). This tool was designed to critically appraise quantitative, qualitative and mixed methods studies and provides a set of criteria and screening questions to provide an overall quality score (24).

### Risk of Bias

Randomized controlled trials will be assessed for risk of bias using the RoB2 (25). Non-randomized comparative studies and pre/post-test intervention studies will be assessed using the ROBINS-1 tool,(26).

### Data synthesis and Presentation

The data will first be assessed for similarity or variation in outcome measures, measurement scales, and types of interventions before it is determined whether a meta-analysis is appropriate.

If sufficient homogeneity exists between the studies, a meta-analysis will be performed, and the findings will be presented graphically in a forest plot to facilitate visual inspection and provide a summary of the data. If a meta-analysis is conducted, heterogeneity will be assessed statistically using the standard χ² and I² tests. Statistical analyses will be performed using a random effects model to facilitate generalization of the findings.

To explore potential sources of heterogeneity, subgroup analyses will be performed based on the intervention’s theoretical basis, including the facilitator, delivery mode, and evaluation tool used to measure effectiveness.

If there is substantial variation in the data and statistical pooling is not feasible, the findings will be presented in a narrative format, supplemented by tables and figures to aid in the interpretation and visualization of the data. The narrative synthesis will be informed by the guidance of the synthesis without meta-analysis in systematic reviews reporting guidelines (SWiM) (27) and will include 1) a detailed description of the methods used for data collection and analysis, such as the types of studies included, the criteria for inclusion and exclusion, and the statistical techniques employed and 2) a comprehensive summary of the findings, highlighting key trends, patterns, and outcomes observed in the data, as well as any limitations or biases identified during the synthesis process.

We will assess the quality of evidence for key outcomes using the Grading of Recommendations Assessment, Development, and Evaluation (GRADE) methodology. This evaluation will encompass several domains, including risk of bias, consistency, directness, precision, and publication bias. Based on these criteria, the evidence will be classified as high, moderate, or low quality.

## Discussion

Several systematic reviews addressing similar aims have previously been carried out (Table 2) however, none of these reviews focused specifically on sexual violence and sexual safety education for adolescents. This systematic review aims to address this gap in sexual health education. Understanding and implementing effective sexual health interventions that address sexual violence and sexual safety that positively influence adolescents’ behaviour, knowledge and skills is crucial. These interventions not only cater to the needs of the adolescents but will also have a broader impact on their families, communities and society. Moreover, they also play a role in alleviating the burden on the healthcare system by promoting sexual safety and addressing issues related to sexual violence. This systematic review will help to identify which educational intervention methods are the most effective, which will inform the development of programs that equip young people with the emotional, cognitive and relational skills required for development of healthy relationships (15).

**Table 1.**
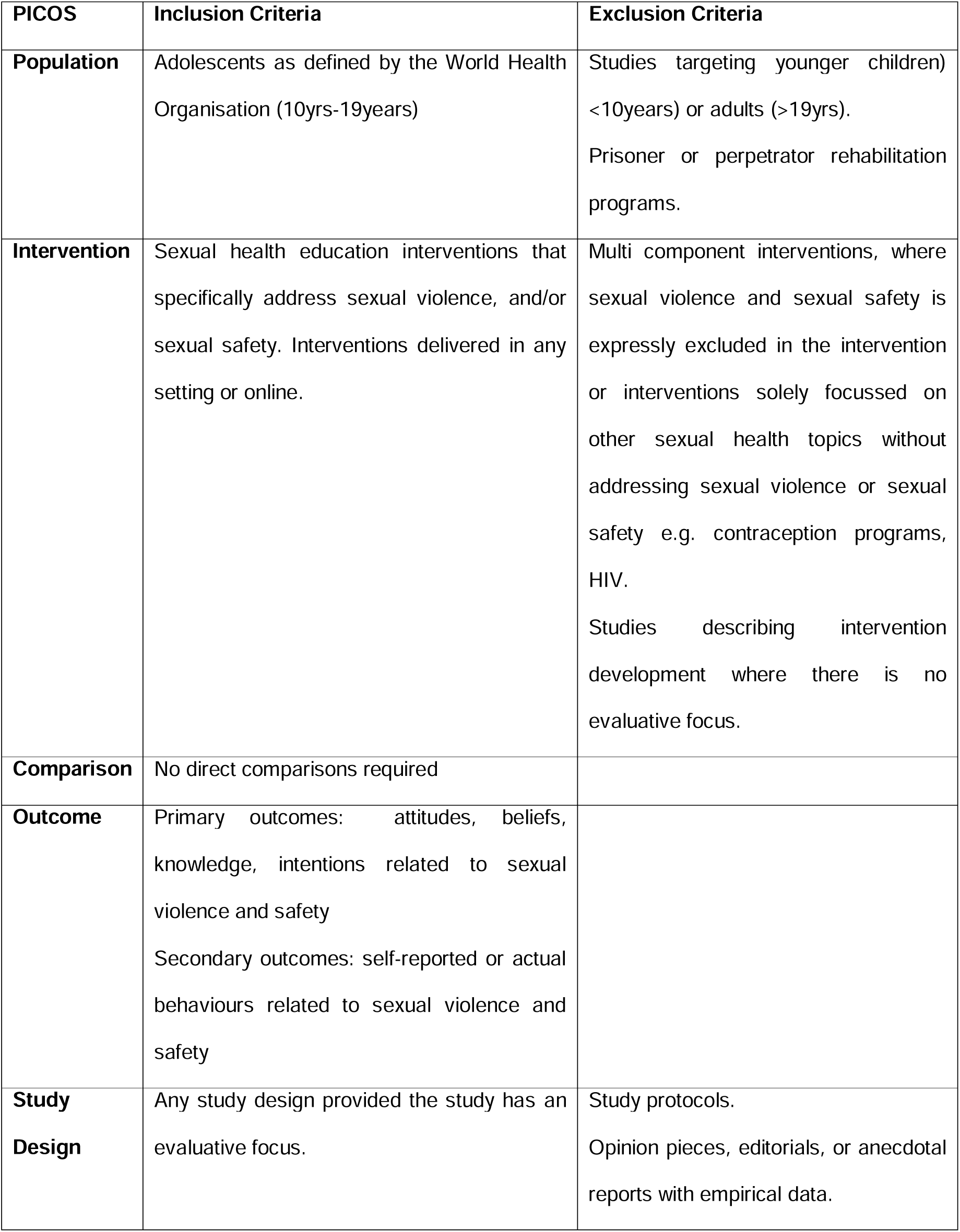
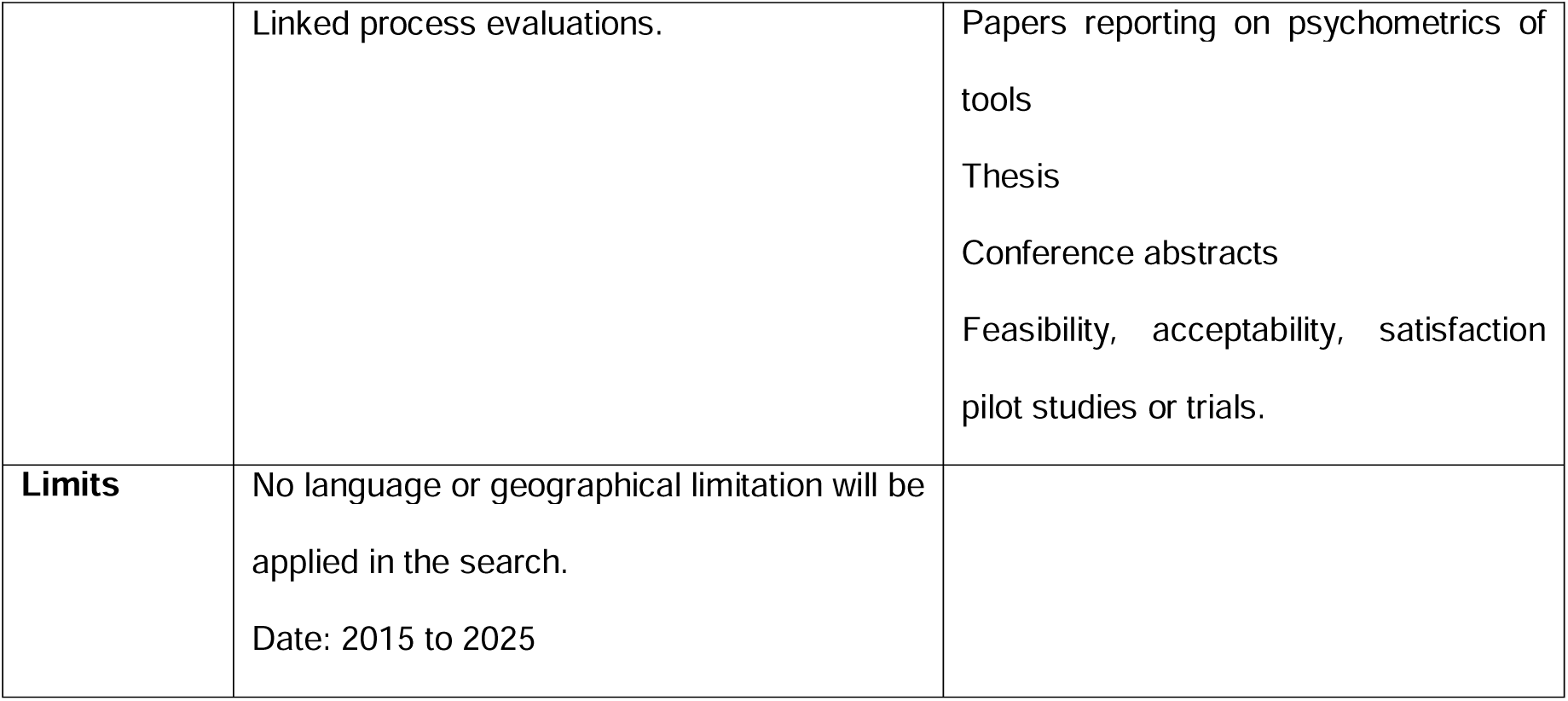
Summary of eligibility criteria.

| **PICOS** | **Inclusion Criteria** | **Exclusion Criteria** |
| --- | --- | --- |
| **Population** | Adolescents as defined by the World Health Organisation (10yrs-19years) | Studies targeting younger children) <10years) or adults (>19yrs).<br>Prisoner or perpetrator rehabilitation programs. |
| **Intervention** | Sexual health education interventions that specifically address sexual violence, and/or sexual safety. Interventions delivered in any setting or online. | Multi component interventions, where sexual violence and sexual safety is expressly excluded in the intervention or interventions solely focussed on other sexual health topics without addressing sexual violence or sexual safety e.g. contraception programs, HIV.<br>Studies describing intervention development where there is no evaluative focus. |
| **Comparison** | No direct comparisons required |  |
| **Outcome** | Primary outcomes: attitudes, beliefs, knowledge, intentions related to sexual violence and safety<br>Secondary outcomes: self-reported or actual behaviours related to sexual violence and safety |  |
| **Study Design** | Any study design provided the study has an evaluative focus. | Study protocols.<br>Opinion pieces, editorials, or anecdotal reports with empirical data. |
|  | Linked process evaluations. | Papers reporting on psychometrics of tools<br>Thesis<br>Conference abstracts<br>Feasibility, acceptability, satisfaction<br>pilot studies or trials. |
| **Limits** | No language or geographical limitation will be applied in the search.<br>Date: 2015 to 2025 |  |

**Table 2.**
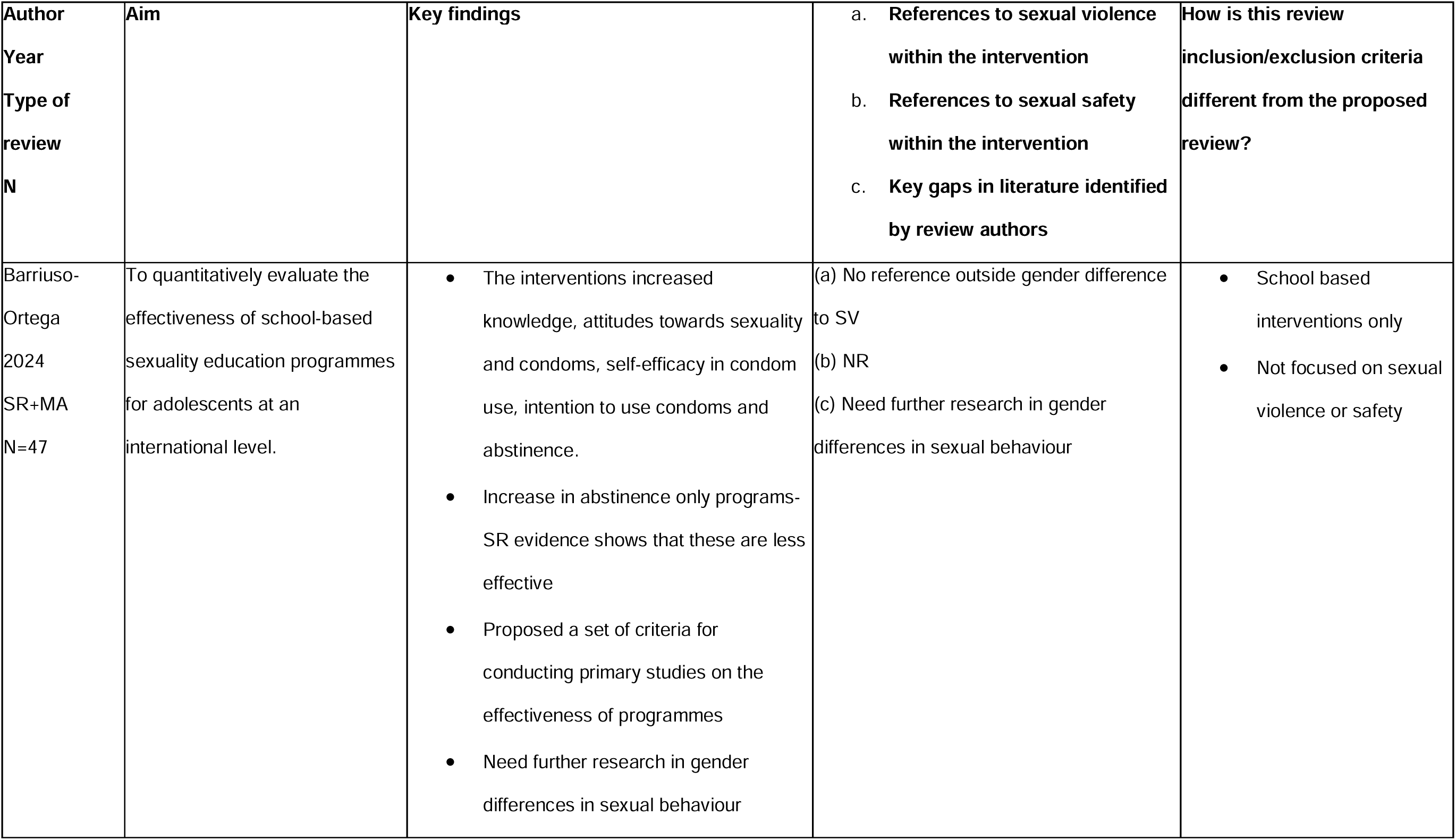
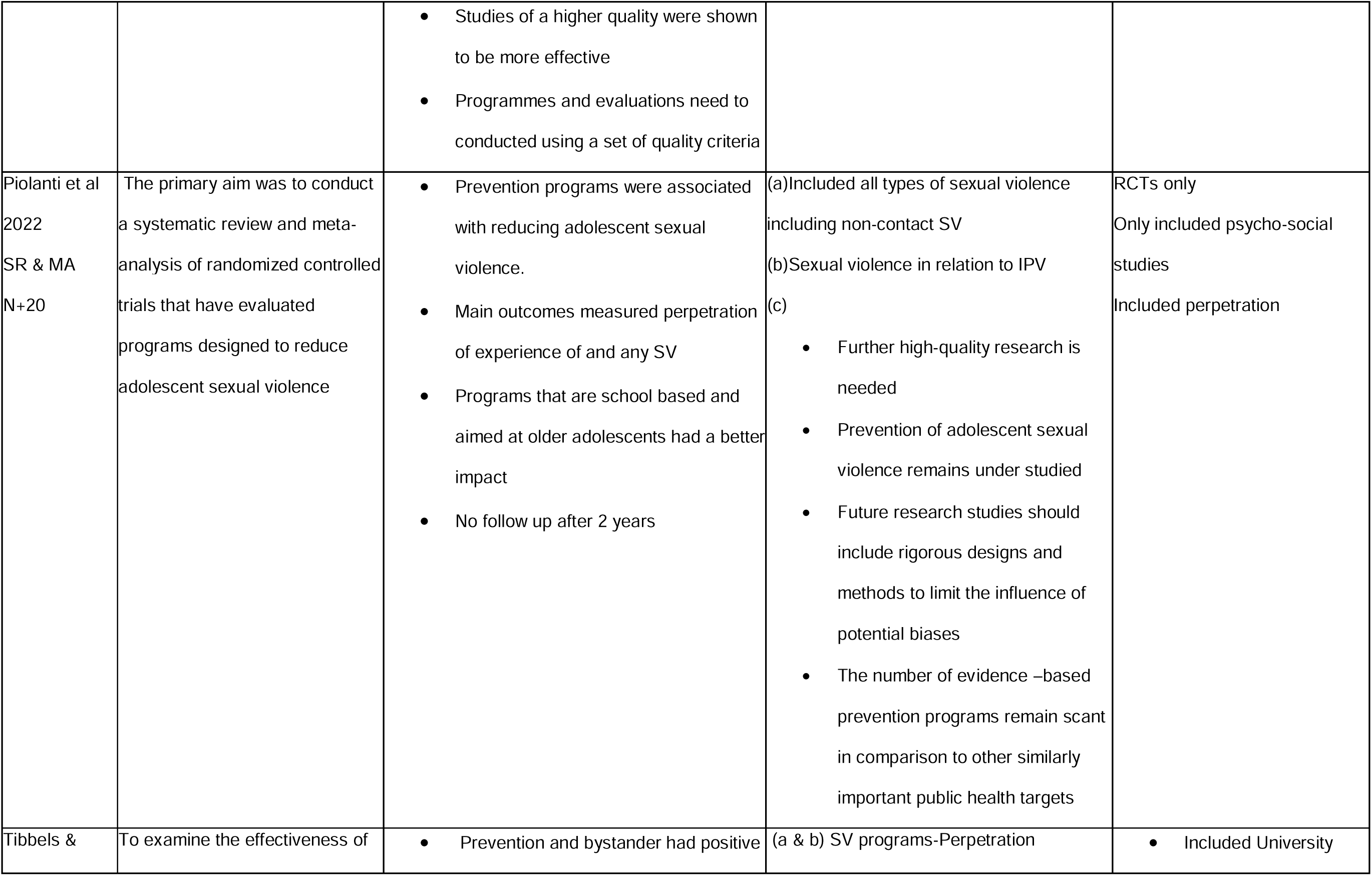
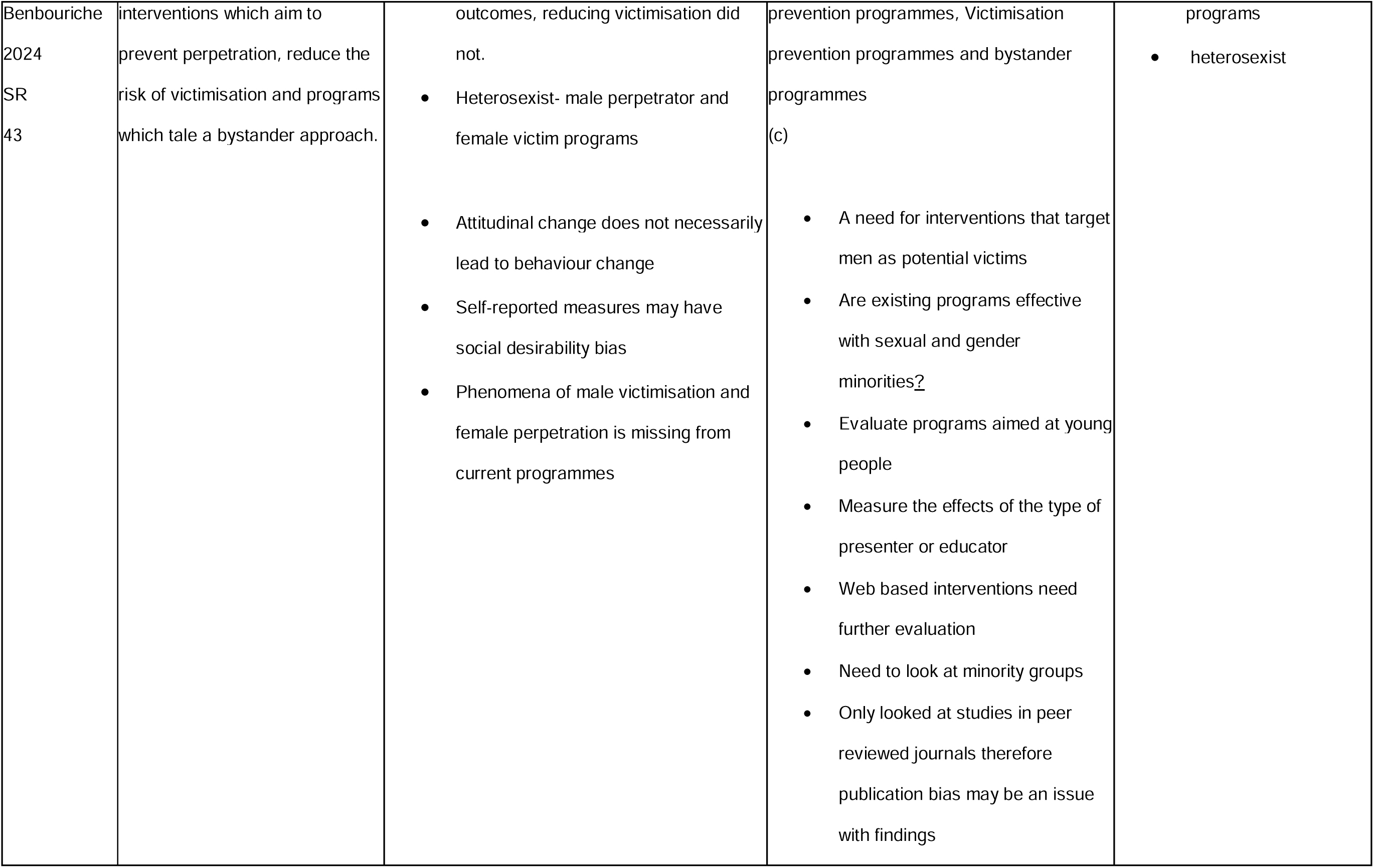
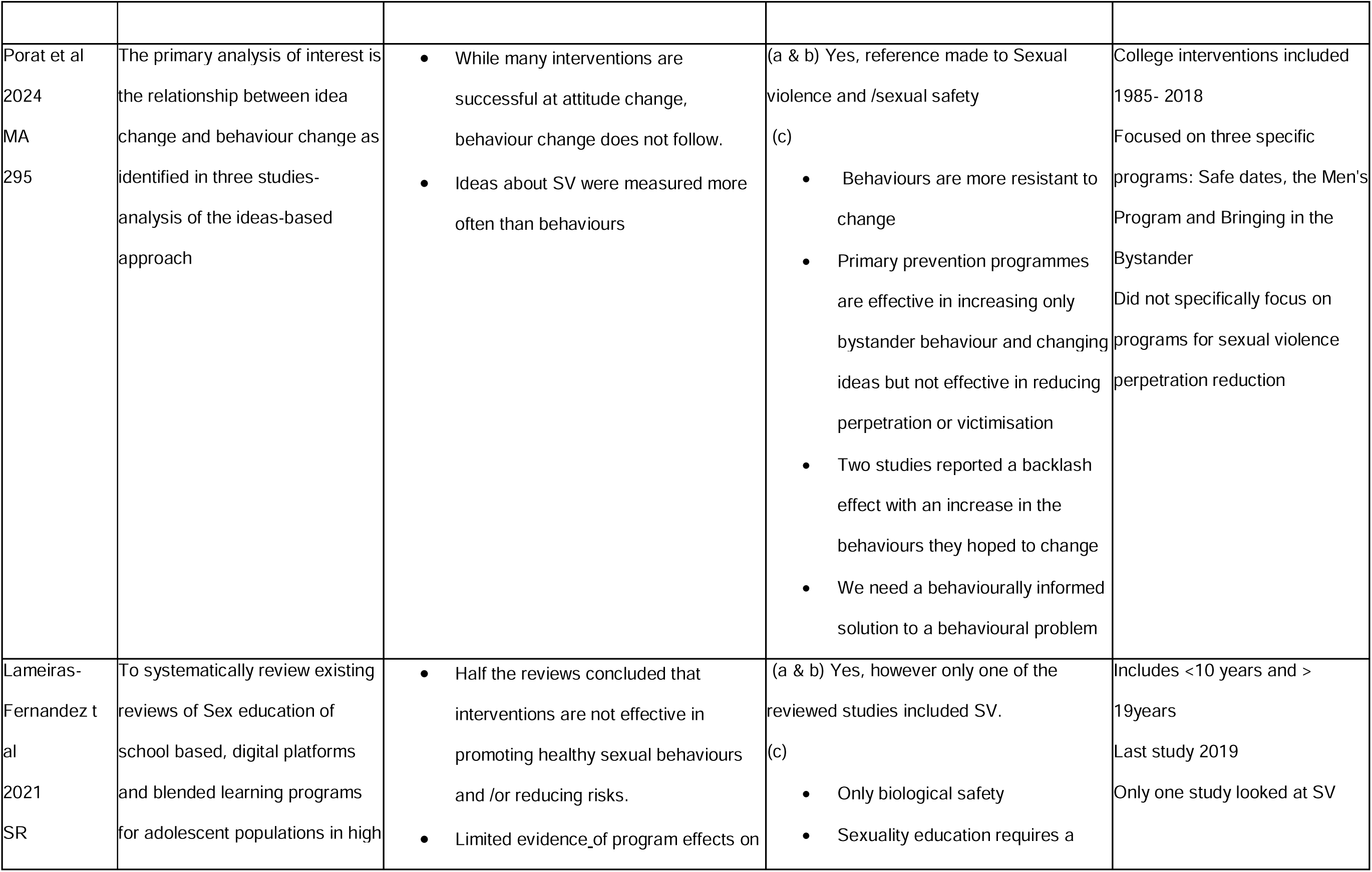
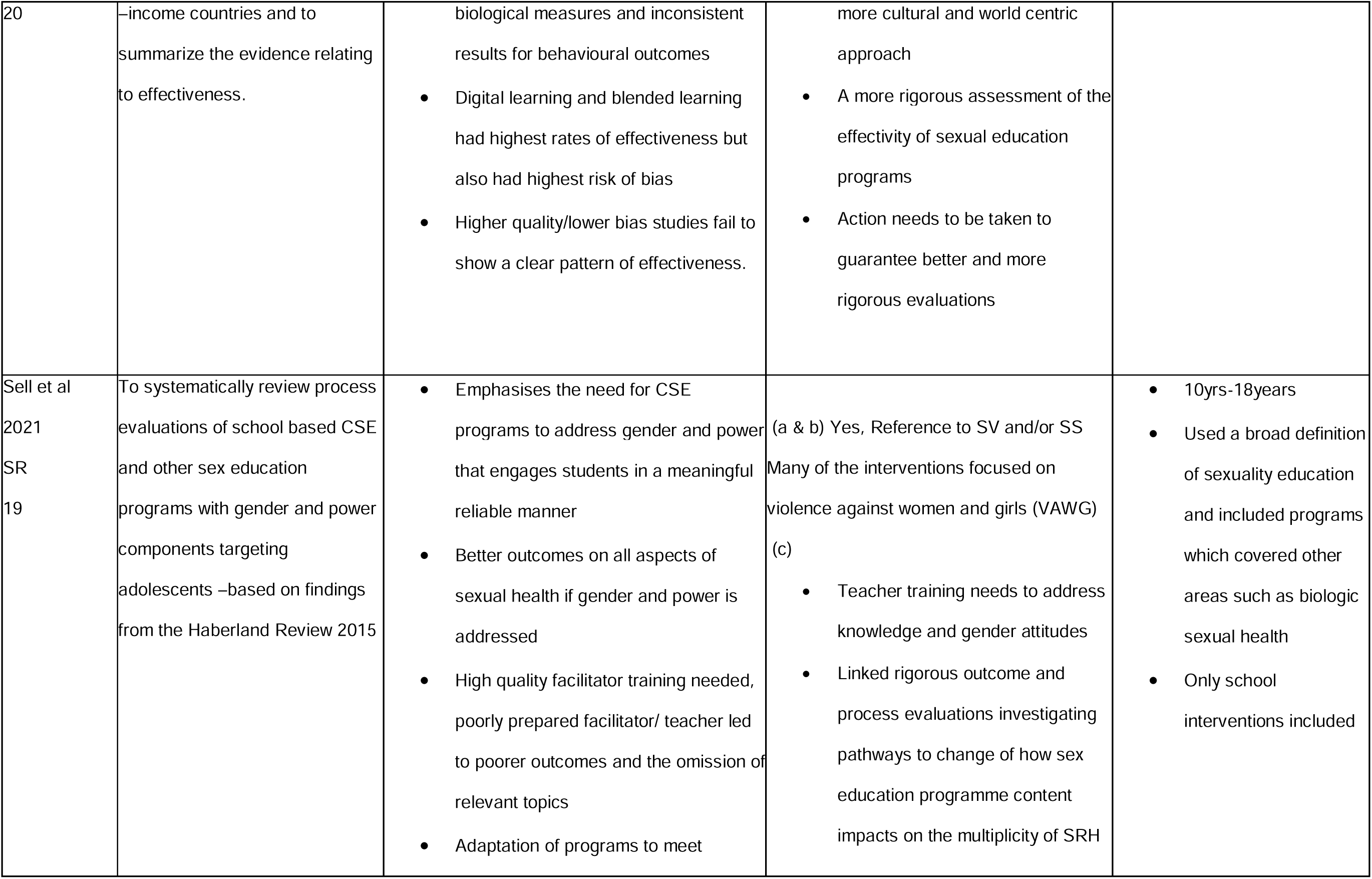
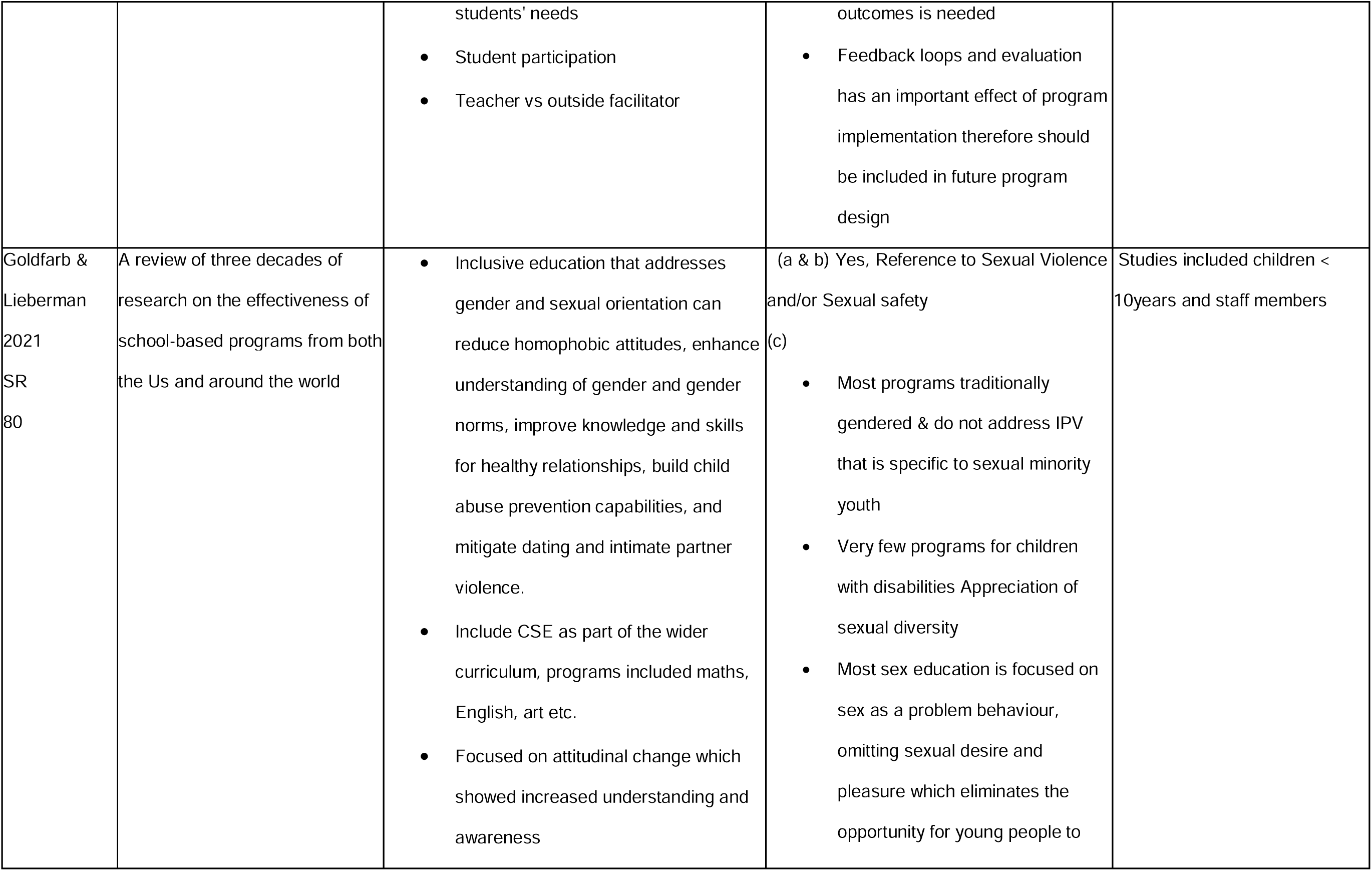
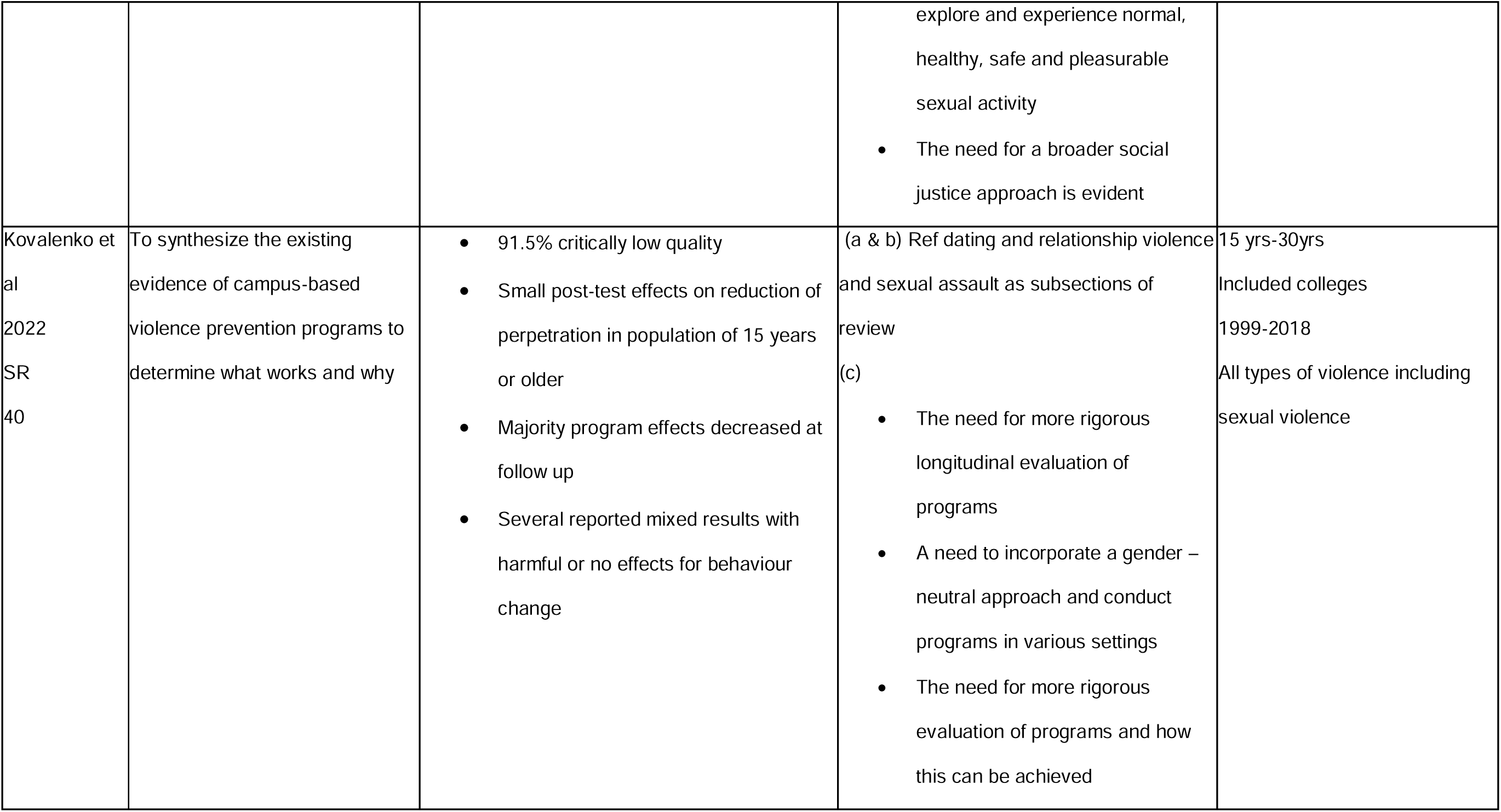
Summary of systematic reviews addressing similar aim.

## Supporting information

Supplement 1 Data Extraction

Supplement 2 Inclusion and exclusion table

Reply to reviewers

Sample 4 Search strategy sample

Supplement 2 Prisma P Checklist

Supplement 5 Table of existing research

## Data Availability

No datasets were generated or analysed during the current study. All relevant data from this study will be made available upon study completion.

## References

1. Ozaki R, Brandon A. Evidence-Based Bystander Programs to Prevent Sexual and Dating Violence in High Schools. Leadership and Research in Education. 2020;5(1):72–97.

2. WHO. Violence Info – Sexual violence. 2022; [cited 2022]; Available from: https://apps.who.int/violence-info/sexual-violence/

3. Banvard-Fox C, Linger M, Paulson DJ, Cottrell L, Davidov DM. Sexual Assault in Adolescents. Prim Care. 2020 Jun;47(2):331–349. doi: 10.1016/j.pop.2020.02.010. Epub 2020 Feb 27. PMID: 32423718; PMCID: PMC7702185.

4. Lameiras-Fernández M, Rodríguez-Castro Y, Carrera-Fernández MV. Effective sex education methods. Int J Environ Res Public Health [Internet]. 2021;18(5). Available from: 10.3390/ijerph18052555

5. Children’s Rights Alliance. A guide on the UN Convention on the Rights of the Child for children and young people. 2010.

6. Council of Europe. Council of Europe Convention on preventing and combating violence against women and domestic violence (Istanbul Convention). Istanbul: Council of Europe; 2014. Available from: https://www.coe.int [Accessed 25 Jan 2025].

7. Council of Europe. Council of Europe Convention on the Protection of Children against Sexual Exploitation and Sexual Abuse (CETS No. 201). Lanzarote; 25 October 2007. Available from: https://www.coe.int/en/web/children/lanzarote-convention.

8. UNESCO. International Guidance on Sexuality Education: An Evidence-Informed Approach. 2018. Available from: https://www.unfpa.org/sites/default/files/pub-pdf/ITGSE.pdf.

9. Caulfield NM, Fergerson AK, Buerke M, Capron DW. Impact of high school sexual education on victimization and rape myth acceptance in college. J Interpers Violence. 2024;1–17. DOI: 10.1177/08862605241257599.

10. Santelli JS, Grilo SA, Choo T, Diaz G, Walsh K, Wall M, Hirsh JS, Wilson PA, Gilbert L, Khan S, Mellins CA. Does pre-college sex education protect against college sexual assault? PLoS ONE. 2018;13(11).

11. Denning DM, Newlands R, Bennet N, Benuto LT. Sexual education experiences and recommendations from survivors. J Aggress Maltreat Trauma. 2022;32(12):1647–1665. DOI: 10.1080/10926771.2022.2146558.

12. Santelli JS, Kantor LM, Grilo SA, Speizer IS, Lindberg LD, Heitel J, et al. Abstinence-Only-Until-Marriage: An updated review of U.S. policies and programs and their impact. J Adolesc Health. 2017;61(3):273–80. doi: 10.1016/j.jadohealth.2017.05.031. PMID: 28842065.

13. Bowring AA, Wright CJC, Douglass C, Gold L, Lim MSC. Features of successful sexual health promotion programs for young people: Findings from a review of systematic reviews. Health Promot J Austr. 2018; 29:46–57.

14. Lamb S, Randazzo R. Effectiveness of a sexual ethics curriculum. J Moral Educ. 2016;45(1):16-

15. UNESCO. The journey towards comprehensive sexuality education: Global status report [Internet]. 2021 [cited 2021 Oct 10]. Available from: https://unesdoc.unesco.org/ark:/48223/pf0000379607

16. Moher D, Shamseer L, Clarke M, Ghersi D, Liberati A, Petticrew M, et al.; PRISMA-P Group. Preferred reporting items for systematic review and meta-analysis protocols (PRISMA-P) 2015 statement. Syst Rev. 2015;4(1):1. doi: 10.1186/2046-4053-4-1. PMID: 25554246; PMCID: PMC4320440.

17. Li T, Higgins JPT, Deeks JJ. Chapter 3: Defining the criteria for including studies and how they will be grouped for the synthesis. In: Higgins JPT, Thomas J, Chandler J, Cumpston M, Li T, Page MJ, Welch VA, editors. Cochrane Handbook for Systematic Reviews of Interventions. Version 6.5 [updated August 2024]. Cochrane; 2024. Available from: https://training.cochrane.org/handbook/current/chapter-03

18. The adolescent health indicators recommended by the Global Action for Measurement of Adolescent health: guidance for monitoring adolescent health at country, regional and global levels. Geneva: World Health Organization; 2024. Licence: CC BY-NC-SA 3.0 IGO.

19. Miele C, Maquigneau A, Joyal CC, Bertsch I, Gangi O, Gonthier H, et al. International guidelines for the prevention of sexual violence: a systematic review and perspective of WHO, UN Women, UNESCO, and UNICEF’s publications. Child Abuse Negl. 2023 Dec;146:106497. doi:10.1016/j.chiabu.2023.106497.

20. Care Quality Commission. Promoting sexual safety through empowerment. February 2020. Available from: [https://office.com?path=‘d627dfb3-a765-4173-ad23-46c14aa3d14a] (https://office.com?path=’d627dfb3-a765-4173-ad23-46c14aa3d14a)’

21. World Health Organization. Comprehensive sexuality education. 2023. Available from: https://www.who.int/news-room/questions-and-answers/item/comprehensive-sexuality-education

22. Page MJ, McKenzie JE, Bossuyt PM, Boutron I, Hoffmann TC, Mulrow CD, et al. The PRISMA 2020 statement: an updated guideline for reporting systematic reviews. PLOS Medicine [Internet]. 2021 [cited 2022 Feb 22];18(3):e1003583 [15p.]. Available from:. [10.1371/journal.pmed.1003583[^i^](https://doi.org/10.1371/journal.pmed.1003 583%5B%5Ei%5E)]

23. Hoffmann TC et al. Better reporting of interventions: TIDieR checklist and guide. BMJ. 2014;348: g1687. doi: 10.1136/bmj.g1687. PMID: 24609605.

24. Hong QN, Pluye P, Fàbregues S, Bartlett G, Boardman F, Cargo M, Dagenais P, Gagnon M-P, Griffiths F, Nicolau B, O’Cathain A, Rousseau M-C, Vedel I. Mixed Methods Appraisal Tool (MMAT), version 2018. Registration of Copyright (#1148552), Canadian Intellectual Property Office, Industry Canada.

25. Sterne JAC, Savović J, Page MJ, Elbers RG, Blencowe NS, Boutron I, et al. RoB 2: a revised tool for assessing risk of bias in randomized trials. BMJ [Internet]. 2019 [cited 2025Apr19];366: l4898.Availablefrom: [10.1136/bmj.l4898[^i^](https://doi.org/10.1136/bmj.l4898% 5B%5Ei%5E)]

26. Sterne JA, Hernán MA, Reeves BC, et al. ROBINS-I: a tool for assessing risk of bias in non-randomised studies of interventions. BMJ. 2016;355: i4919. doi:10.1136/bmj.i4919. PMID:27733354; PMCID: PMC5062054.

27. Campbell M, McKenzie JE, Sowden A, Katikireddi SV, Brennan SE, Ellis S, et al. Synthesis Without Meta-analysis (SWiM) in systematic reviews: reporting guideline. BMJ [Internet]. 2020 [cited 2025 Apr 19];368:l6890. Available from: [10.1136/bmj.l6890[^i^](https://doi.org/10.1136/bmj.l6890%5B%5Ei%5E)].

28. Mullinax, M., Mathur, S., Santelli, J. (2017). Adolescent Sexual Health and Sexuality Education. In: Cherry, A., Baltag, V., Dillon, M. (eds) International Handbook on Adolescent Health and Development. Springer, Cham. 10.1007/978-3-319-40743-2_8

29. Barriuso S, Fernandez-Hawrylak M, Heras-Sevilla D. Sex education in adolescence: A systematic review of programmes and meta-analysis. Children Youth Serv Rev. 2024; 166:107926. doi:10.1016/j.childyouth.2024.107926.

30. Tibbels S, Benbouriche M. Sexual Violence in Young People: A Systematic Literature Review of Prevention Programmes. Sex Cult. 2024;28(1):1–24. doi:10.1007/s12119-023-10187-8.

31. Porat R, Gantman A, Green SA, Pezzuto JH, Paluck EL. Preventing Sexual Violence: A Behavioral Problem Without a Behaviorally Informed Solution. Psychol Sci Public Interest. 2024 May;25(1):4–29. doi: 10.1177/15291006231221978. PMID: 38832574; PMCID: PMC11151714.

32. Kovalenko AG, Abraham C, Graham-Rowe E, Levine M, O’Dwyer S. What Works in Violence Prevention Among Young People? A Systematic Review of Reviews. Trauma Violence Abuse. 2022 Dec;23(5):1388–1404. doi: 10.1177/1524838020939130. Epub 2020 Jul 17. PMID: 32677554; PMCID: PMC9606003.

33. Piolanti A, Jouriles EN, Foran HM. Assessment of Psychosocial Programs to Prevent Sexual Violence During Adolescence: A Systematic Review and Meta-analysis. JAMA Netw Open. 2022;5(11): e2240895. doi:10.1001/jamanetworkopen.2022.40895

