## Supplement 1 Data Extraction for "“Effectiveness of Sexual Health Education Interventions Addressing Sexual Violence and Sexual Safety for Adolescents: A Systematic Review Protocol”"

**Intervention Characteristics (using TIDieR Framework)**

| **Author**  **Year**  **Country** | **Name and Objective of Intervention** | **Study Setting** | **Characteristics of participants** | **Mode of Delivery & Duration** | **Facilitators of intervention & training** | **Intervention Tailored** | **Modifications to intervention** | **Was the program/intervention evaluated** | **Fidelity maintained** | **Limitations** |
| --- | --- | --- | --- | --- | --- | --- | --- | --- | --- | --- |

**Evaluation Characteristics**

| **Author/study** | **Mode of evaluation** | **Characteristics of evaluators** | **Full intervention or component of a larger intervention** | **Timing of Evaluation(s) and no. of participants** | **Primary Outcome Measure** | **Secondary Outcome measure (IF applicable)** | **Pre-tested validated tools utilized** | **Main Findings** | **Other** |
| --- | --- | --- | --- | --- | --- | --- | --- | --- | --- |

or practices

**Quality Assessment of Included Articles**

| **Author/study** | **Study Design** | **Selection Bias** | **Consistency** | **Blinding** | **Directness** | **Precision** | **Publication Bias** | **Additional Notes/other** |
| --- | --- | --- | --- | --- | --- | --- | --- | --- |

Note: 1=Strong, 2=moderate, 3=Weak, N/R+ not recorded

**Summary of Findings Table**

| **Outcome** | **Number of Studies** | **Participants** | **Effect (95%CI)** | **Quality of Evidence (GRADE)** | **MMAT Criterion met** | **Recommendations for future research, policy or practice** | **Other** |
| --- | --- | --- | --- | --- | --- | --- | --- |
| Behaviour |  |  |  |  |  |  |  |
| Skills |  |  |  |  |  |  |  |
| Knowledge |  |  |  |  |  |  |  |
| Attitude |  |  |  |  |  |  |  |
| Intent to change |  |  |  |  |  |  |  |

Behaviour: Impact on behavioural outcomes as part of the intervention

Skills: Enhanced understanding or retention of information

Attitude: Shifts in attitude or perspectives resulting from the intervention

Intent to change: Evidence of willingness or plans to modify behaviour or practices
