## Supplement 2 Inclusion and exclusion table for "“Effectiveness of Sexual Health Education Interventions Addressing Sexual Violence and Sexual Safety for Adolescents: A Systematic Review Protocol”"

Inclusion and Exclusion Criteria

|  | Description, inclusion | Exclusion Criteria |
| --- | --- | --- |
| Population | Adolescents as defined by the World Health Organisation (10yrs-19years) | Studies targeting younger children) <10years) or adults (>19yrs).  Prisoner or perpetrator rehabilitation programs. |
| Intervention | Sexual health education interventions that specifically address sexual violence and/or sexual safety. Interventions delivered in any setting or online. | Multi-component interventions, where sexual violence and sexual safety are expressly excluded in the intervention or interventions solely focussed on other sexual health topics without addressing sexual violence or sexual safety, e.g., contraception programs, HIV.  Studies describing intervention development where there is no evaluative focus. |
| **Comparison** | No direct comparisons are required. |  |
| **Outcome** | Primary outcomes: attitudes, beliefs, knowledge, and intentions related to sexual violence and safety  Secondary outcomes: self-reported or actual behaviours related to sexual violence and safety |  |
| **Study Design** | Any study design provided the study has an evaluative focus.  Linked process evaluations. | Study protocols.  Opinion pieces, editorials, or anecdotal reports with empirical data.  Papers reporting on psychometrics of tools  Thesis  Conference abstracts  Feasibility, acceptability, satisfaction pilot studies or trials. |
| **Setting** | No geographical or language limitation |  |
| Time Limits | 2015-2025 |  |
