## Supplementary material for "“Effectiveness of Sexual Health Education Interventions Addressing Sexual Violence and Sexual Safety for Adolescents: A Systematic Review Protocol”": Reply to reviewers

Dear Reviewers,

Many thanks for your feedback on our submission: **"Effectiveness of Sexual Health Education Interventions Addressing Sexual Violence and Sexual Safety for Adolescents: A Systematic Review Protocol."**

The following additions and amendments have been made to enhance our submission:

- **Inclusion and Exclusion Criteria**: Detailed on pages 4 and 5, with a summary table on page 6.

- **Data Extraction Sheet**: Included as a supplement.

- **Flowchart for Inclusion/Exclusion Decisions**: A PRISMA chart has been added to the supplement.

- **Outcome Assessments**: The manuscript has incorporated a GRADE amendment.

Thank you for allowing us to improve our protocol. If you need any further information or details, please do not hesitate to contact me.

| **Recommendation** | **Page** |  |
| --- | --- | --- |
| Defined Search Criteria | Added as a supplement |  |
| Inclusion and Exclusion Criteria | Page 4 & 5 with a summary table on page 6 |  |
| Data Extraction Sheet | Supplement |  |
| Flowchart showing how inclusion/exclusion decisions were made | PRISMA Chart included as a supplement |  |
| Outcome Assessments | GRADE | An amendment added to the manuscript |
