## Supplementary material for "“Effectiveness of Sexual Health Education Interventions Addressing Sexual Violence and Sexual Safety for Adolescents: A Systematic Review Protocol”": Sample 4 Search strategy sample

| **Cinahl Search Strategy Outline** | **Search Terms (CINAHL Title & Abstract)** |
| --- | --- |
| **PICO** |  |
| **Population:** | “young people” OR adolescen* OR teen* OR youth OR “school student” OR “young person” OR students |
| **Intervention No.1** | “sex ed” OR “sex education” OR “CSE” OR program* OR intervention OR “relationship education” OR “prevention education” OR “sex* health education” OR “school health education” OR educat* OR learn* OR “consent education” OR “consent program” OR “healthy relationships education” OR “sexual safety” OR “sexual safety education” |
| **Context** | “sexual consent” OR “sexual assault” OR “sexual violence” OR IPV OR “dating violence” OR “dating abuse” OR “sexual violence prevention” OR “sexual harassment” OR “sexual exploitation” |
| **Comparison** | No Comparator |
| **Outcomes** | Skill* OR attitude* OR belief* OR knowledge OR behaviour OR behavior OR “attitude to sexual violence” OR “intent to change” OR understanding OR “safety outcomes” Or “self-efficacy” OR empowerment OR “program evaluation” OR effectiveness OR impact OR outcomes OR efficacy OR evalu* |
| **Limiters** | Language: Any  Age group: Adolescents (10-19 years)  Publication type: Peer-reviewed, empirical studies  Date range: 2015-2025 |
