## Supplement 5 Table of existing research for "“Effectiveness of Sexual Health Education Interventions Addressing Sexual Violence and Sexual Safety for Adolescents: A Systematic Review Protocol”"

Table X Summary of systematic reviews addressing similar aim

| **Author**  **Year**  **Type of review**  **N** | **Aim** | **Key findings** | a. **References to sexual violence within the intervention** b. **References to sexual safety within the intervention** c. **Key gaps in literature identified by review authors** | **How is this review inclusion/exclusion criteria different from the proposed review?** |
| --- | --- | --- | --- | --- |
| Barriuso-Ortega  2024  SR+MA  N=47 | To quantitatively evaluate the effectiveness of school-based sexuality education programmes for adolescents at an international level. | - The interventions increased knowledge, attitudes towards sexuality and condoms, self-efficacy in condom use, intention to use condoms and abstinence. - Increase in abstinence only programs-SR evidence shows that these are less effective - Proposed a set of criteria for conducting primary studies on the effectiveness of programmes - Need further research in gender differences in sexual behaviour - Studies of a higher quality were shown to be more effective - Programmes and evaluations need to conducted using a set of quality criteria | - No reference outside gender difference to SV - No reference to sexual safety - Need further research in gender differences in sexual behaviour | - School based interventions only - Not focused on sexual violence or safety |
| Piolanti et al  2022  SR & MA  N+20 | The primary aim was to conduct a systematic review and meta-analysis of randomized controlled trials that have evaluated programs designed to reduce adolescent sexual violence | - Prevention programs were associated with reducing adolescent sexual violence. - Main outcomes measured perpetration of, experience of and any SV - Programs that are school based and aimed at older adolescents had a better impact - No follow up after 2 years | - Included all types of sexual violence including non-contact SV - Sexual violence in relation to IPV - Further high-quality research is needed - Prevention of adolescent sexual violence remains under studied - Future research studies should include rigorous designs and methods to limit the influence of potential biases - The number of evidence –based prevention programs remain scant in comparison to other similarly important public health targets | RCTs only  Only included psycho-social studies  Included perpetration |
| Tibbels & Benbouriche  2024  SR  43 | To examine the effectiveness of interventions which aim to prevent perpetration, reduce the risk of victimisation and programs which tale a bystander approach. | - Prevention and bystander had positive outcomes, reducing victimisation did not. - Heterosexist- male perpetrator and female victim programs - Attitudinal change does not necessarily lead to behaviour change - Self-reported measures may have social desirability bias - Phenomena of male victimisation and female perpetration is missing from current programmes | - SV programs-Perpetration prevention programmes, Victimisation prevention programmes and bystander programmes - Yes - A need for interventions that target men as potential victims - Are existing programs effective with sexual and gender minorities? - Evaluate programs aimed at young people - Measure the effects of the type of presenter or educator - Web based interventions need further evaluation - Need to look at minority groups - Only looked at studies in peer reviewed journals therefore publication bias may be an issue with findings | - Included University programs   heterosexist |
| Porat et al  2024  MA  295 | The primary analysis of interest is the relationship between idea change and behaviour change as identified in three studies-analysis of the ideas-based approach | - While many interventions are successful at attitude change, behaviour change does not follow. - Ideas about SV were measured more often than behaviours | - Yes - Yes - Behaviours are more resistant to change - Primary prevention programmes are effective in increasing only bystander behaviour and changing ideas but not effective in reducing perpetration or victimisation - Two studies reported a backlash effect with an increase in the behaviours they hoped to change - We need a behaviourally informed solution to a behavioural problem | College interventions included  1985- 2018  Focused on three specific programs: Safe dates, the Men's Program and Bringing in the Bystander  Did not specifically focus on programs for sexual violence perpetration reduction |
| Lameiras-Fernandez t al  2021  SR  20 | To systematically review existing reviews of Sex education of school based, digital platforms and blended learning programs for adolescent populations in high –income countries and to summarize the evidence relating to effectiveness. | - Half the reviews concluded that interventions are not effective in promoting healthy sexual behaviours and /or reducing risks. - Limited evidence of program effects on biological measures and inconsistent results for behavioural outcomes - Digital learning and blended learning had highest rates of effectiveness but also had highest risk of bias - Higher quality/lower bias studies fail to show a clear pattern of effectiveness. | - Yes, however only one of the reviewed studies. - Only biological safety - Sexuality education requires a more cultural and world centric approach - A more rigorous assessment of the effectivity of sexual education programs - Action needs to be taken to guarantee better and more rigorous evaluations | Includes <10 years and > 19years  Last study 2019  Only one study looked at SV |
| Sell et al  2021  SR  19 | To systematically review process evaluations of school based CSE and other sex education programs with gender and power components targeting adolescents –based on findings from the Haberland Review 2015 | - Emphasises the need for CSE programs to address gender and power that engages students in a meaningful reliable manner - Better outcomes on all aspects of sexual health if gender and power is addressed - High quality facilitator training needed, poorly prepared facilitator/ teacher led to poorer outcomes and the omission of relevant topics - Adaptation of programs to meet students' needs - Student participation - Teacher vs outside facilitator | - Yes - Yes - Many of the interventions focused on violence against women and girls (VAWG) - Teacher training needs to address knowledge and gender attitudes - Linked rigorous outcome and process evaluations investigating pathways to change of how sex education programme content impacts on the multiplicity of SRH outcomes is needed - Feedback loops and evaluation has an important effect of program implementation therefore should be included in future program design | - 10yrs-18years - Used a broad definition of sexuality education and included programs which covered other areas such as biologic sexual health - Only school interventions included |
| Goldfarb & Lieberman  2021  SR  80 | A review of three decades of research on the effectiveness of school-based programs from both the Us and around the world | - Inclusive education that addresses gender and sexual orientation can reduce homophobic attitudes, enhance understanding of gender and gender norms, improve knowledge and skills for healthy relationships, build child abuse prevention capabilities, and mitigate dating and intimate partner violence. - Include CSE as part of the wider curriculum, programs included maths, English, art etc. - Focused on attitudinal change which showed increased understanding and awareness | - Yes - Yes - Most programs traditionally gendered & do not address IPV that is specific to sexual minority youth - Very few programs for children with disabilities Appreciation of sexual diversity - Most sex education is focused on sex as a problem behaviour, omitting sexual desire and pleasure which eliminates the opportunity for young people to explore and experience normal, healthy, safe and pleasurable sexual activity - The need for a broader social justice approach is evident | Studies included children < 10years and staff members |
| Kovalenko et al  2022  SR  40 | To synthesize the existing evidence of campus-based violence prevention programs to determine what works and why | - 91.5% critically low quality - Small post-test effects on reduction of perpetration in population of 15 years or older - Majority program effects decreased at follow up - Several reported mixed results with harmful or no effects for behaviour change | - Ref dating and relationship violence and sexual assault as subsections of review - The need for more rigorous longitudinal evaluation of programs - A need to incorporate a gender –neutral approach and conduct programs in various settings - The need for more rigorous evaluation of programs and how this can be achieved | 15 yrs-30yrs  Included colleges  1999-2018  All types of violence including sexual violence |

Completed on 03.03.2025
